# SARS-CoV-2 Reinfection is Preceded by Unique Biomarkers and Related to Initial Infection Timing and Severity: an N3C RECOVER EHR-Based Cohort Study

**DOI:** 10.1101/2023.01.03.22284042

**Authors:** Emily Hadley, Yun Jae Yoo, Saaya Patel, Andrea Zhou, Bryan Laraway, Rachel Wong, Alexander Preiss, Rob Chew, Hannah Davis, Christopher G Chute, Emily R Pfaff, Johanna Loomba, Melissa Haendel, Elaine Hill, Richard Moffitt, the N3C and RECOVER consortia, the N3C and RECOVER consortia

## Abstract

Although the COVID-19 pandemic has persisted for over 2 years, reinfections with SARS-CoV-2 are not well understood. We use the electronic health record (EHR)-based study cohort from the National COVID Cohort Collaborative (N3C) as part of the NIH Researching COVID to Enhance Recovery (RECOVER) Initiative to characterize reinfection, understand development of Long COVID after reinfection, and compare severity of reinfection with initial infection. We validate previous findings of reinfection incidence (5.9%), the occurrence of most reinfections during the Omicron epoch, and evidence of multiple reinfections. We present novel findings that Long COVID diagnoses occur closer to the index date for infection or reinfection in the Omicron BA epoch. We report lower albumin levels leading up to reinfection and a statistically significant association of severity between first infection and reinfection (chi-squared value: 9446.2, p-value: 0) with a medium effect size (Cramer’s V: 0.18, DoF = 4).

## INTRODUCTION

Throughout the COVID-19 pandemic, hundreds of millions of SARS-CoV-2 cases have been confirmed worldwide.^1^ However, an infection with SARS-CoV-2 does not confer lasting immunity, particularly in the context of immunologic escape displayed by new variants.^2^ Reports of SARS-CoV-2 reinfection are well documented, and whole genome sequencing analysis has confirmed reinfections from SARS-CoV-2 variants that are genetically distinct from an initial SARS-CoV-2 infection.^3^ Reinfections are concerning because they may interfere with the development of herd immunity.^4^

In this work, we seek to contribute to the growing literature on SARS-CoV-2 reinfections with novel findings from a large cohort of more than 1.5 million individuals in the electronic health record (EHR)-based N3C Data Enclave. We first characterize reinfection by describing incidence, attributes, and biomarkers. We then explore the severity of reinfection as measured by hospitalization and consider the relationship of reinfection and Long COVID. Finally, we discuss findings and suggest opportunities for further research.

Incidence estimates of reinfections among persons who experienced a SARS-CoV-2 infection are low, ranging from 0.2% to 5.5%.^5,6^ A review of laboratory studies found that the time from primary SARS-CoV-2 infection to reinfection can range from 19 to 293 days.^7^ Guidelines generally suggest that a new positive COVID-19 antigen or PCR test should be considered a reinfection if it occurred at least 60 to 90 days after initial infection.^5,6,8–11^ A few studies document cases of two or three infections, noting that third infections were mainly associated with the transmission of the Omicron variant.^10,11^ In this work, we aim to validate these findings with analyses from a larger cohort.

Biomarkers are an important tool for characterizing a disease. Existing research has explored the relationship of severity of COVID-19 with biomarkers such as laboratory indicators of inflammation, dysregulated coagulation and end-organ dysfunction.^12–14^ Although studies of reinfection have been less common, one study that characterized patients with suspected reinfection showed increased rates of metabolic failure and similar rates of renal and hepatic failure with reinfection compared to their index encounter, but did not further analyze these findings using laboratory biomarkers.^13^ Most studies of biomarkers related to COVID-19 infection are limited to the time period during infection, with limited insight into the trajectories of laboratory indicators between infection and reinfection. In this work, we aim to use biomarkers captured in EHR data between the index date and reinfection to add to the characterization of the disease in reinfection and population-level trends in biomarkers.

Considerable interest exists regarding the severity of reinfection as compared to initial infection. Hospitalization can be an indicator of disease severity because more severe disease often requires treatment. Studies looking at reinfection and hospitalization have generally found that rates of hospitalization following reinfection were similar to or lower than rates of hospitalization following initial SARS-CoV-2 infection.^15,16^ One study found that the reduced risk of hospitalization following reinfection persisted when disaggregated by age.^17^ No study has clearly disaggregated by severity of hospitalization, such as considering the distinction between an emergency department (ED) visit, an inpatient hospitalization, and an inpatient hospitalization requiring intensive care. In this work, we aim to assess the association between the severity of initial infection and severity of first reinfection to contribute more granular findings disaggregated by severity of hospitalization.

Less attention has been given to the relationship of reinfections to post-acute sequelae of SARS-CoV-2 infection (PASC) or Long COVID.^18^ PASC is understood as complications resulting from SARS-CoV-2 that persist or occur de novo for at least 4 weeks post-infection, and Long COVID is the clinical diagnosis for these conditions. Long COVID is associated with commonly reported symptoms including fatigue that interferes with daily life, fever, cough, sleep problems, difficulty breathing, and difficulty thinking.^19^ Existing work suggests that reinfection can increase the risk of post-acute sequelae in the pulmonary organ systems.^20^ Additional knowledge about the relationship of reinfections and Long COVID could help inform interested parties who may be concerned that reinfections could contribute to the incidence of Long COVID. In this work, we contribute novel findings on the relationship of reinfection and Long COVID.

## METHODS

This study uses individual EHR data stored in the N3C Data Enclave as part of the NIH Researching COVID to Enhance Recover (RECOVER) Initiative. The RECOVER Initiative seeks to understand, treat, and prevent PASC. For more information on RECOVER, visit https://recovercovid.org. The N3C Data Enclave provides access to harmonized EHRs from more than 75 health sites with data from over 16 million patients.^21,22^ We used N3C data from version 87 (8/2/2022), which has 63 contributing sites, for the current investigation. The N3C Data Enclave’s Palantir Foundry platform (2021, Denver, CO), a secure analytics platform, was used for data access and analysis.

### Key Definitions

We describe the following key definitions for the study cohort, reinfection, COVID-19 variant epochs, and Long COVID.

#### Study Cohort Definition, Inclusion, and Exclusion Criteria

The study inclusion criteria include (1) having an International Classification of Diseases-10-Clinical Modification (ICD-10) COVID-19 diagnosis code (U07.1) or a positive SARS-CoV-2 PCR or antigen test between March 1, 2020, and May 1, 2022; the earliest of these events was considered the COVID-19 index date; (2) reinfection events (if any) occurring before July 1, 2022; (3) being 18 years of age or older; (4) having at least one recorded healthcare visit in the two years prior to index; (5) having at least one recorded healthcare visit more than 60 days after the COVID-19 index date; (6) being from a hospital partner with data that has been updated in the last two months prior to July 1, 2022; (7) being from a hospital partner with at minimum 100 hospitalizations related to a first known COVID-19 infection and at minimum 25 hospitalizations related to a COVID-19 reinfection. A total of 1,597,490 individuals met these criteria.

#### Definition of Reinfection

A COVID-19 reinfection was defined as a positive SARS-CoV-2 PCR or antigen test that occurred 60 or more days after a COVID-19 infection index date. The date of the test was considered the first COVID-19 reinfection index date. Subsequent reinfections were defined as a new positive SARS-CoV-2 PCR or antigen test that occurred 60 or more days after each reinfection index date. Although a threshold of 90 days for reinfection post index date is common in the literature, other findings suggest that nearly all patients stop shedding SARS-CoV-2 within 60 days of infection and many stop shedding much sooner than that.^23–26^ Based on these findings and support from the RECOVER clinician advisory panel, 60 days was selected as a more appropriate threshold.

#### Definition of the COVID-19 Variant Epoch

We define the following COVID-19 variant epochs based on the patient’s COVID-19 diagnosis code (U07.1) or a positive SARS-CoV-2 PCR or antigen test date: Wild-type COVID-19 (March 01, 2020-November 30, 2020), Alpha/Beta/Gamma variant (December 01, 2020-May 31, 2021), Delta variant (June 01, 2021-October 31, 2022), Omicron variant (November 01, 2021-March 11, 2022), and Omicron BA variant (March 12, 2022-August 01, 2022).^27^

#### Definition of Long COVID

Patients with a Long COVID diagnosis were identified with the U09.9 or B94.8 ICD-10-CM diagnosis codes. The U09.9 code was implemented in October 2021 for providers to use in a clinical setting with patients experiencing ongoing conditions after a COVID-19 infection, commonly understood as Long COVID. Many hospital sites appear to have rapidly adopted the use of U09.9 once it became available.^28^ The B94.8 code is not specific to COVID-19 and instead represents sequelae of other specified infectious and parasitic diseases. This code was rarely used prior to the pandemic, but it started seeing considerably more use in November 2020. The use of this code is understood to represent Long COVID diagnoses prior to the availability of U09.9.^28^ For the purposes of the Long COVID analysis, we limited the study cohort to individuals at sites that had at least 250 uses of either the U09.9 code after October 1, 2021 or the B94.8 code after November 1, 2020. Eligible reinfections for U09.9 or B94.8 had to occur after the respective dates of use of the codes. This subcohort included 1,568,810 individuals.

### Statistical Analysis

In this work, we perform three main analyses focused on describing characterizing reinfection, understanding reinfection severity, and exploring the relationship of reinfection and long COVID. All analysis and visualization were done in the N3C enclave using SQL, Python (v3.6), and R (v3.6), including ggplot2, survival, and survminer packages.

#### Characterization of Reinfection

We used two approaches to characterize reinfection. The first is a cohort summary where we calculate summary statistics related to reinfection and disaggregate by demographic characteristics. Chi-square tests were used for categorical variables and Student’s t-test or ANOVA were used for continuous variables (**Table 1, Supplementary Table 2, 3**). Time to reinfection analysis was based on Kaplan-Meier curves from the *survival* package in R. This analysis was performed using the date of the COVID-19 index date (the date of earliest diagnosis or positive test) and the date of event (first reinfection date) as endpoints.

**Table 1:** Descriptive characteristics of reinfected and non-reinfected COVID-19 positive patients.

| Category | Variable | No Reinfection<br>(N=1505855) | Reinfected<br>Patients<br>(N=91635) | Total<br>(N=1597490) | P value |
| --- | --- | --- | --- | --- | --- |
| **Age,**<br>**Mean (SD)** | Age | 49.14(18.26) | 44.31(17.71) | 48.85(18.27) | <0.0001 |
| **Sex**<br>**(N, %)** | Female | 946166 (62.83) | 61989 (67.65) | 1008155 (63.11) |  |
|  | Male | 559280 (37.14) | 29617 (32.32) | 588897 (36.86) | <0.0001 |
|  | No sex<br>Information | 409 (0.03) | 29 (0.03) | 438 (0.03) |  |
| **Race (N, %)** | White | 1141563 (75.81) | 69852 (76.23) | 1211415 (75.83) |  |
|  | Black | 199891 (13.27) | 12871 (14.05) | 212762 (13.32) |  |
|  | Asian | 24615 (1.63) | 1022 (1.12) | 25637 (1.6) |  |
|  | Others | 33996 (2.26) | 2222 (2.42) | 36218 (2.27) | <0.0001 |
|  | No race<br>Information | 105790 (7.03) | 5668 (6.19) | 111458 (6.98) |  |
| **Ethnicity**<br>**(N, %)** | Not<br>Hispanic | 1197960 (79.55) | 73706 (80.43) | 1271666 (79.6) |  |
|  | Hispanic | 106956 (7.1) | 7925 (8.65) | 114881 (7.19) | <0.0001 |
|  | No<br>Ethnicity<br>information | 200939 (13.34) | 10004 (10.92) | 210943 (13.2) |  |

**Table 2:** Comparison of Severity of First and Reinfection with SARS-CoV-2.

| Count<br>Row (R)%<br>Column (C)%<br>Table (T)%<br>among those<br>with a<br>reinfection* | Severity of Reinfection |  |  |  |  |  | Total |
| --- | --- | --- | --- | --- | --- | --- | --- |
| Severity of Initial Infection | No Documented Reinfection | Mild with No ED Visit or Hospitalization Around Reinfection Index | Mild with ED Visit Around Reinfection Index | Moderate with Hospitalization Around Reinfection Index | Severe with ECMO or IMV or Vasopressor During Hospitalization Around Reinfection Index | Death Within 60 Days of Reinfection Index |  |
| Mild with No ED Visit or Hospitalization Around COVID Index | 1288792 | 71126<br>R: 88.8%<br>C: 90.9%<br>T: 77.6% | 4970<br>R: 6.2%<br>C: 67.5%<br>T: 5.4% | 2961<br>R: 3.7%<br>C: 65.3%<br>T: 3.2% | 414<br>R: 0.5%<br>C: 64.7%<br>T: 0.4% | 622<br>R: 0.8%<br>C: 72.0%<br>T: 0.7% | 1368885<br>(80093 multiple infections) |
| Mild with ED Visit Around COVID Index | 115740 | 4357<br>R: 64.9%<br>C: 5.6%<br>T: 4.8% | 1820<br>R: 27.1%<br>C: 24.7%<br>T: 2.0% | 432<br>R: 6.4%<br>C: 9.5%<br>T: 0.5% | 67<br>R: 1.0%<br>C: 10.5%<br>T: <0.1% | 39<br>R: 0.6%<br>C: 4.5%<br>T: <0.1% | 122455<br>(6715 multiple infections) |
| Moderate with Hospitalization Around COVID Index | 85959 | 2496<br>R: 57.3%<br>C: 3.2%<br>T: 2.7% | 523<br>R: 12.0%<br>C: 7.1%<br>T: 0.6% | 1034<br>R: 23.8%<br>C: 22.8%<br>T: 1.1% | 124<br>R: 2.9%<br>C: 19.4%<br>T: 0.1% | 176<br>R: 4.0%<br>C: 20.4%<br>T: 0.2% | 90312<br>(4353 multiple infections) |
| Severe with ECMO or IMV or Vasopressor During Hospitalization Around COVID Index | 9357 | 251<br>R: 52.9%<br>C: 0.3%<br>T: 0.2% | 52<br>R: 12.0%<br>C: 0.7%<br>T: <0.1% | 109<br>R: 23.0%<br>C: 2.4%<br>T: 0.1% | 35<br>R: 7.4%<br>C: 5.5%<br>T: <0.1% | 27<br>R: 5.7%<br>C: 3.1%<br>T: <0.1% | 9831<br>(474 multiple infections) |
| Death Within 60 Days of COVID Index | 6007 | N/A | N/A | N/A | N/A | N/A | 6007 |
| **Total** | 1505855 | 78230 | 7365 | 4536 | 640 | 864 | 1597490 |
| *The shaded region indicates the values used for calculation of row, column, and table percentages among those with a SARS-CoV-2 reinfection. Individuals without a reinfection are not included in this calculation. |  |  |  |  |  |  |  |

The second approach to characterizing reinfection is with biomarkers. We explored the trajectories of various biomarkers around COVID-19 initial and subsequent index dates from patients with and without a reinfection. Biomarker measurements included laboratory values of ferritin, fibrinogen, C-reactive protein, procalcitonin, white blood cell count, absolute lymphocyte count, absolute neutrophil count, erythrocyte sedimentation rate, albumin, D-dimer, alanine transaminase (ALT), aspartate transaminase (AST), and serum creatinine. Units were harmonized across data partners, and clinically infeasible values were excluded according to standard N3C data quality protocols.^22^ Measurements were taken from 100 days prior to and 180 days after the COVID-19 index date in patients with and without reinfection. The same time frame was used for collection around the first reinfection index date in individuals with at least one reinfection. Laboratory values were reported separately for hospitalized and nonhospitalized patients. The median laboratory value of each biomarker, with upper (75%) and lower (25%) quartiles, was binned by 7-day intervals and visualized according to time from COVID-19 infection or reinfection index date. For patients with more than one measurement of the same laboratory test in a day, values were averaged.

#### Severity of Reinfection

Severity of infection was measured with records of COVID-associated hospitalization, which was defined as an inpatient visit with a start date 1 day prior to 16 days after the COVID-19 index date with a COVID diagnosis code used during the visit. A COVID-associated ED visit was defined as an ED visit with a start date 1 day prior to 16 days after the COVID-19 index date. These thresholds were intended to capture hospitalizations and ED visits that are related to COVID. Severity of infection is assessed with hospitalization metrics. Four levels of severity are considered: mild infection that does not require an ED visit or hospitalization, mild infection that requires an ED visit, moderate infection that requires hospitalization, and severe infection that requires hospitalization and use of extracorporeal membrane oxygenation (ECMO), invasive mechanical ventilation (IMV), or vasopressors. Vasopressors were included in addition to the more intensive ECMO and IMV because some hospitals may be limited in their ability to provide ECMO or IMV. We compared the severity of the first COVID-19 infection versus severity of first reinfection using a pivot table with row, column, and table percentages along with a chi-square test for association. Death after initial infection or reinfection is also included in the table.

#### Reinfections and Long COVID

The subcohort of individuals described in Section 2.1.4 was used for analysis of reinfections and Long COVID. Kaplan-Meier curves were calculated to explore the differences in time to Long COVID diagnosis following initial infection versus reinfection. Time to event analysis was performed using the initial COVID-19 index date and with the first reinfection index date (for those with one or more reinfections) to the first B94.8 or U09.9 diagnosis.

## RESULTS

The study cohort included 1,597,490 adults (age: mean 48.85 years, standard deviations (SD) 18.27; 63.11% female) (**Table 1, Supplementary Table 1**). The study cohort contains data from 38 health facility data partners. Women make up nearly two-thirds of the study cohort and a larger proportion of individuals with reinfections. The skew in sex can be attributed to the inclusion requirement for at least two visits prior to COVID diagnosis (**Supplementary Figure 1**). The sex ratio of individuals hospitalized with COVID-19 was more balanced (**Supplementary Table 2**).

**Figure 1.**
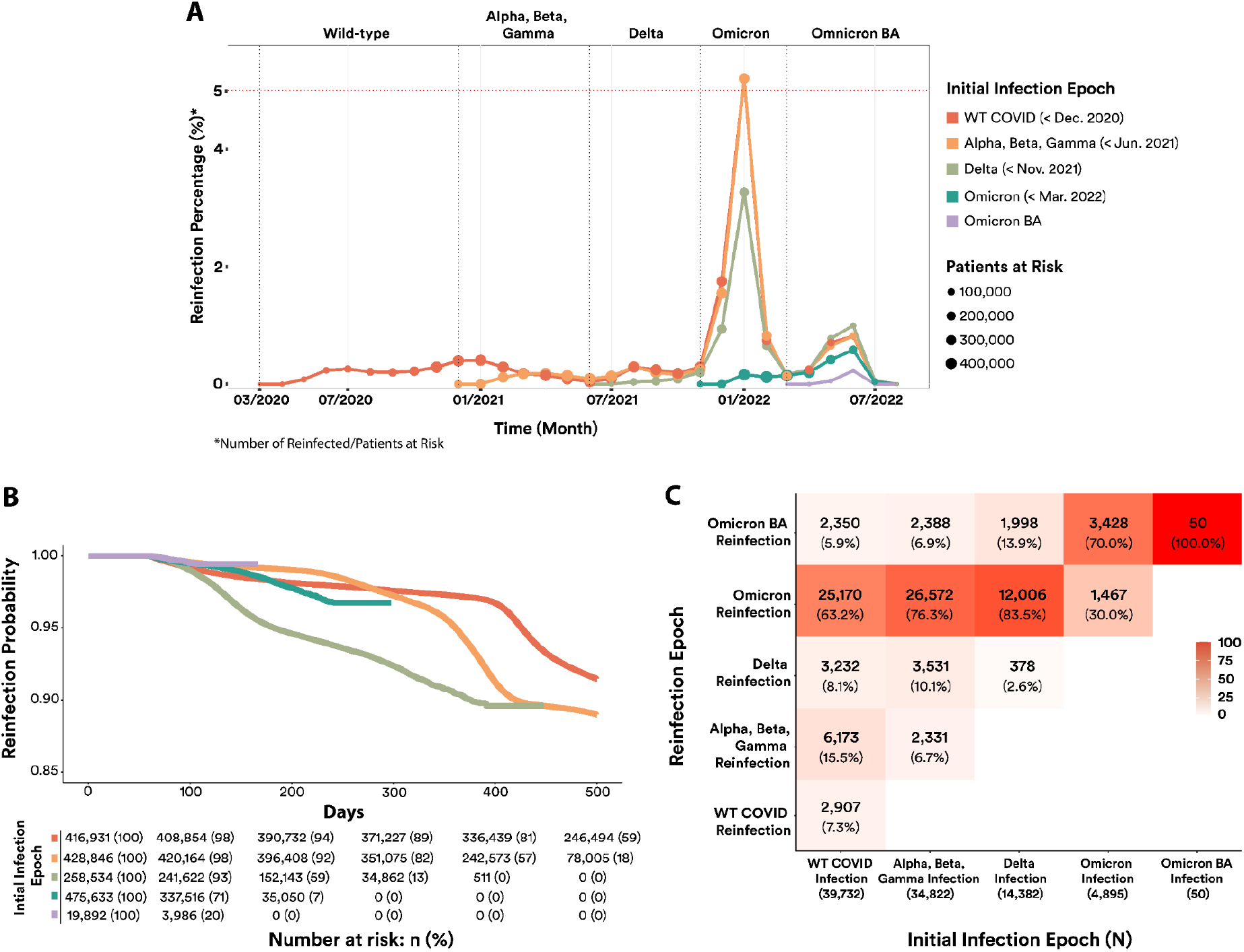

### Characterization of Reinfection

**Table 1** describes the study cohort and highlights differences between the subgroup with no reinfections and the subgroup with at least one reinfection. A total of 5.9% of the study cohort had at least one documented reinfection. A documented reinfection was defined as a positive SARS-CoV-2 PCR or antigen test that occurred 60 or more days after a COVID-19 infection index date. Home COVID-19 tests administered outside a healthcare setting were not included in the dataset. The subgroup of reinfected patients tends to be younger and more likely to have documented race and ethnicity information. Although the majority of individuals (N=89,357) had only one reinfection, a small group (N=102) had three or more reinfections. This group was characterized by non-Hispanic, male, White race, and older age groups compared to other groups (**Supplementary Table 1**). Supplementary Table 1 shows Table 1 further disaggregated by number of reinfections.

**Figure 1** illustrates three approaches for understanding the occurrence of reinfection as it relates to COVID-19 variants. **Figure 1A** shows the percentage of patients at risk that had a reinfection in a given month. Distinct colors are used to indicate the epoch of initial infection and the size of the dot illustrates the number of persons with a reinfection. This figure is useful for appreciating the varying likelihood of reinfection while accounting for individuals who passed away following their first infection since these individuals are no longer considered at risk. This figure shows the largest increase in reinfections in the Omicron epoch among individuals with initial infections during the WT COVID and Alpha, Beta, Gamma periods which overlap in the Omicron epoch, and a smaller increase among those first infected in the Delta epoch. The difference between these variants is smaller for reinfections in the Omicron BA epoch.

**Figure 1B** is a KM-curve that shows time to event between initial COVID-19 infection and first COVID-19 reinfection by COVID-19 variant. This figure is useful for understanding the days to infection, including demonstrating that in many cases, reinfection occurred more than 100 days after the initial COVID-19 infection. **Figure 1B** also shows that this analysis is not particularly sensitive to the decision to use a 60-day threshold rather than a 90-day threshold because few reinfections occur in the 60- to 90-day window for any variant. We recognize that this figure cannot appropriately account for individuals who passed away following their first infection; because of this violation of proportional hazards, we have chosen to not report odds ratios because they may be misinterpreted. **Figure 1C** most clearly details the relationship between the variant of initial infection and the variant of reinfection, highlighting that reinfections in the Omicron time period were particularly common among individuals initially infected in the Wild-type COVID-19 and the Alpha, Beta, and Gamma epochs.

We evaluated biomarker trends in **Figure 2**, comparing the median laboratory values of patients with and without a reinfection. Comparisons were made between the index date of initial infection and subsequent reinfection date. Biomarkers of hepatic inflammation (ALT and AST) were less elevated during acute reinfection compared to initial COVID-19 infection and normalized over a similar time period. However, albumin trends show that among patients with reinfection, albumin levels were persistently lower after initial COVID-19 infection and that levels were also lower prior to the reinfection date.

**Figure 2.**
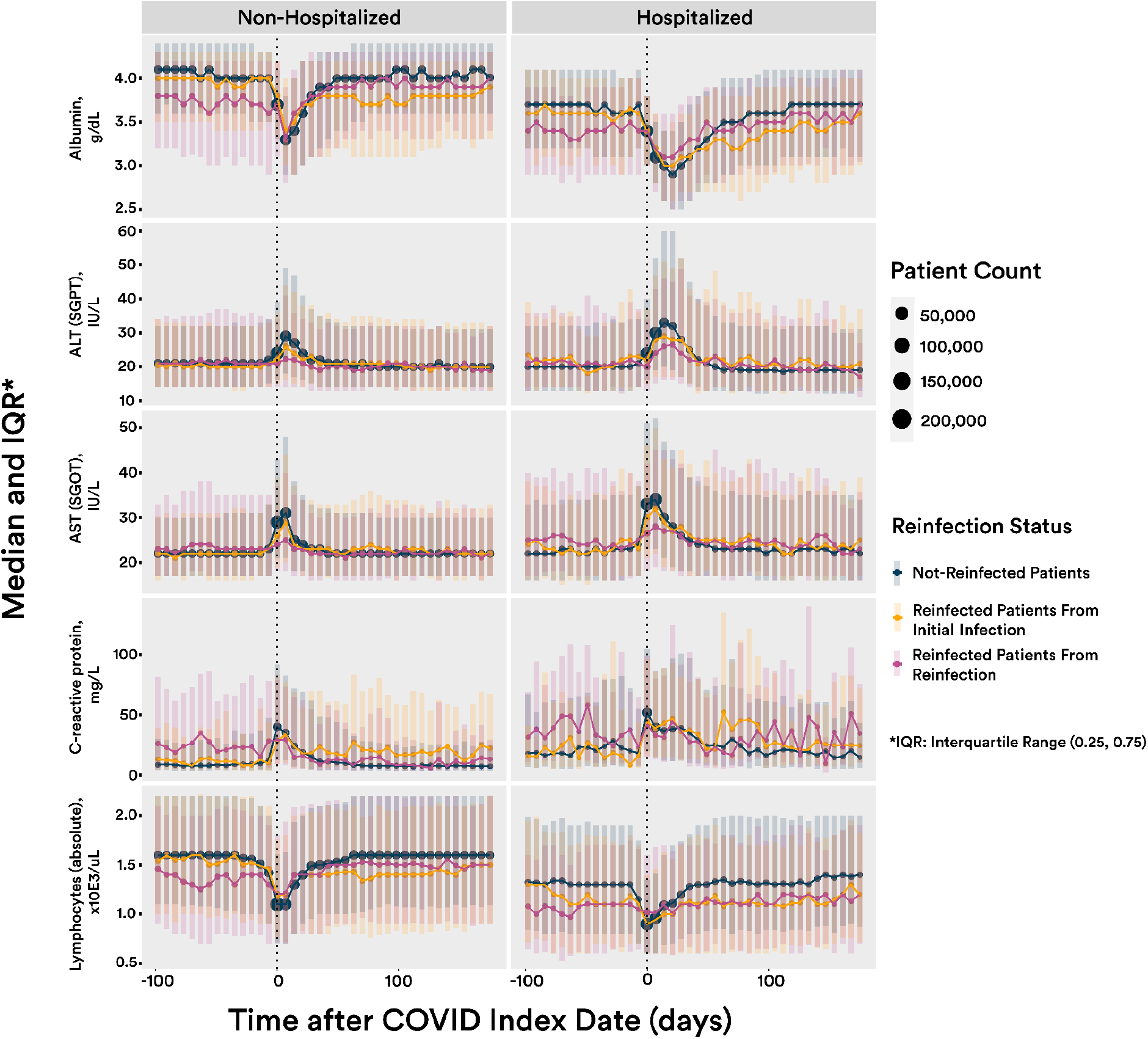

### Severity of Reinfection

We explored the characteristics of severity of infection and hospitalization in **Table 2**. The “No Documented Reinfection” column highlights that the large majority of individuals, even those with ED visits or hospitalization, do not have documentation of a reinfection. The shaded portion of Table 2 summarizes results from individuals with a documented reinfection. We performed a chi-square test of independence for both the entirety of Table 2 (with “No Reinfection” included) and for the shaded portion of Table 2. For the entire table, we found a statistically significant difference (chi-squared value: 9049.2, p-value: 0), although with a negligible effect size (Cramer’s V: 0.04, DoF = 4). For the shaded portion of the table, we found a statistically significant difference (chi-squared value: 9446.2, p-value: 0) with a medium effect size (Cramer’s V: 0.18, DoF = 4). These results, particularly those among individuals who experience reinfection, suggest that the severity of reinfection may not be independent from the severity of initial infection.

Among individuals with a documented reinfection, the majority (77.6%) experienced both mild first and reinfections that did not require an ED visit or hospitalization. A smaller percentage of individuals (12.6%) required either an ED visit or hospitalization or passed away following their reinfection as compared to individuals who required an ED visit or hospitalization or passed away after first infection (14.4%). Among those with either an ED visit or hospitalization during their first infection, a substantial proportion also required an ED visit or hospitalization or passed away following their first reinfection. This included nearly half (47.1%) of those who experienced a severe first infection.

Among individuals who experienced a severe initial infection, 7.4% experienced a severe reinfection and another 5.7% passed away following reinfection. More individuals with a severe initial infection (23.0%) had a moderate reinfection which required hospitalization but did not require use of ECMO or IMV. The largest proportion of individuals who experienced a severe reinfection had mild initial infections with no ED visit or hospitalization (64.7%). Individuals with mild initial infections also made up the majority of those with a mild infection requiring an ED visit or moderate hospitalization for reinfection. The second largest group of individuals with a mild reinfection requiring an ED visit was those who also had a first infection that was mild and required an ED visit (27.1%). A similar proportion of individuals with a moderate initial infection requiring hospitalization (23.8%) also required hospitalization during reinfection.

We provide additional detail in Supplementary Table 2 with a breakdown of demographic details for patients who experience hospitalization for COVID-19. In Supplementary Table 3, we provide Table 2 disaggregated by age, noting that disclosure requirements may limit the robustness of conclusions.

### Reinfection and Diagnosis of Long COVID

Finally, **Figure 3** shows the Kaplan-Meier curves for time to Long COVID diagnosis after initial infection versus first reinfection. For most variant epochs, incidence of new Long COVID diagnoses after reinfection is lower than after initial COVID-19 infection. The exception is Omicron BA where incidence of new Long COVID diagnoses after reinfection in Omicron BA is higher than after initial COVID-19 in Omicron BA. The rate of Long COVID diagnoses following the first infection was largest in the Delta epoch and smallest in the Omicron BA epoch. This contrasts with the incidence of Long COVID diagnoses after first reinfection which is largest in the Omicron BA epoch and smallest following reinfection during the Delta epoch.

**Figure 3.**
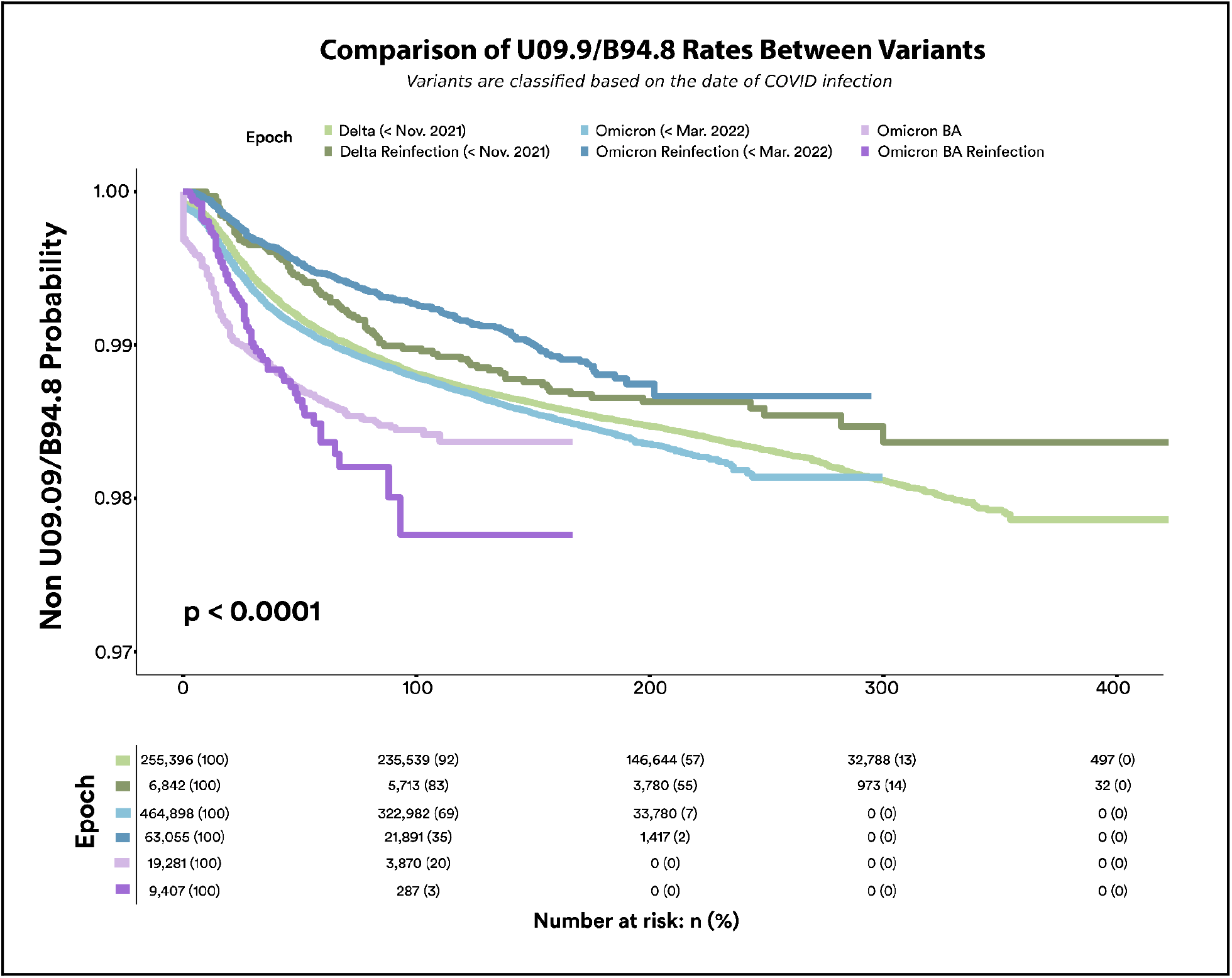

## DISCUSSION

The overall proportion of individuals with at least one documented reinfection in the cohort (5.7%) was slightly larger than the upper bound of incidence of reinfection noted in the literature (5.5%).^5^ This is likely an underestimate given the advent of home testing, particularly in more recent epochs when home testing has been even more accessible. Similar to existing findings, the large majority of reinfections occurred during the Omicron epoch. Figure 1 suggests that this finding holds regardless of the epoch of the initial COVID-19 infection. The large number of reinfections during Omicron makes it challenging to draw conclusions about comparing reinfections between variants because there may be other factors, such as adherence to mask or social distance policies, that impacted likelihood of exposure and subsequent reinfections.

However, it is notable that in Figure 1, initial infection during Delta appeared to be more protective against reinfection during Omicron than initial infection during WT COVID or Alpha, Beta, Gamma. This difference disappeared during Omicron BA. Previous studies have documented reinfections up to 293 days after initial COVID-19 infection.^7^ This study finds evidence of reinfections that occur more than 300 days and up to 500 days after initial COVID-19 infection. This work also validates the occurrence of multiple reinfections, including a small subset of individuals with three or more reinfections.

The mean age of reinfected individuals is close to 5 years lower than the mean age of those without reinfection. This aligns with literature that suggests that reinfections are more commonly reported for younger individuals: one study found that the reinfection rate was highest among those aged 18 to 29 years old^29^ while public health data collected from September 2021 through September 2022 in the state of Washington found disproportionately large numbers of individuals with reinfections in the 18- to 34- and 35- to 49-year old age groups.^30^ One possible explanation is that younger age groups are less likely to use COVID-19 preventative measures like social distancing and more likely to engage with others for work and leisure, leading to multiple exposures and reinfections.^31^ Another possible explanation is that younger individuals are less likely to be vaccinated and may be more susceptible to reinfection.^32^ A third possible explanation is that older individuals were more likely to die following initial infection, so more reinfections occurred among younger individuals.^33^ A fourth possible explanation is that young adults have high rates of asymptomatic and paucisymptomatic infection which may be less protective of reinfection.^34^ More research is needed to explore these potential explanations.

Women make up nearly two-thirds of the study cohort and a larger proportion of those with a documented reinfection. Some studies have also noted larger proportions of women than men with reinfections and suggested that there may be relevant differences in immune response by sex.^35–37^ However, Supplemental Figure 1 suggests that the inclusion criteria requiring at least two visits prior to COVID-19 index date contributes to this imbalance. Previous research also suggests that women are more likely to utilize health services and demonstrate health-seeking behavior.^38,39^ The sex imbalance in these findings may be more likely associated with differential healthcare utilization rather than biological differences.

We explore the characterization of reinfection with biomarkers. Biomarkers have been well studied in acute COVID-19 infection and several laboratory markers have been associated with higher severity of infection and mortality.^40,41^ Our work extends knowledge of biomarkers to reinfections. In finding that biomarkers of hepatic inflammation were less elevated during acute reinfection compared to acute initial infection, we contribute a novel result that has not yet been reported. Another new finding is that albumin appears to be lower leading up to reinfection. Previous studies have suggested that hypoalbuminemia is common in COVID-19 patients and dynamic monitoring of serum albumin can be a useful tool for evaluating the prognosis of COVID-19 infections. We suggest that further work may explore if lower albumin levels may be a predictor of COVID-19 reinfection.

Similar to previous studies measuring severity of reinfection through hospitalization, we find that most individuals did not require an ED visit or hospitalization for either first infection or reinfection. We also find that the total proportion of individuals requiring an ED visit or hospitalization or passing away is similar between first infection and reinfection. However, when we further interrogate these results, we contribute novel findings that individuals who were hospitalized for an acute initial infection are potentially at much greater risk for hospitalization during reinfection. The effect size is medium when we consider the results among those with reinfections, noting that we cannot account for what would have happened among those who passed away following their first infection.

We first consider the varying degrees of severity of initial infection among individuals who also have a reinfection. Nearly two thirds of individuals who had a mild initial infection that did include an ED visit (64.9%) did not require an ED visit or hospitalization or pass away after reinfection, but a substantial proportion (27.1%) recorded an ED visit for their reinfection. This proportion is larger than the respective proportion of individuals who had a moderate or severe hospitalization during first infection and an ED visit following reinfection. Further analysis could investigate if the group of individuals who visited an ED for both first and reinfection reflects ED-seeking behavior that may result from convenience of the ED, limited access to other healthcare options, or health insurance status.^42^

More than a quarter of individuals with either a moderate or severe first infection requiring hospitalization also experienced hospitalization for reinfection. Although this is concerning, a promising finding is that the proportion experiencing a severe reinfection requiring ECMO, IMV, or vasopressors during hospitalization was small (2.9% of those with moderate first infection and 7.4% of those with severe first infection). One possible explanation is that experiencing hospitalization during reinfection may be related to experiencing hospitalization during first infection, but the hospitalization may be less severe. Another possible explanation is that clinical thresholds for this treatment or clinical behavior may have changed over time. We also observe that the proportion of individuals who pass away following reinfection is higher than the proportion who experience a severe reinfection among those with an initial moderate infection and similar for those who have a severe initial infection. We suggest further analysis to better understand attributes, such as comorbidities, that are predictive of moderate and severe reinfections.

This study contributes novel findings of the relationship of reinfection with Long COVID diagnosis. The largest proportion of Long COVID diagnoses occur among individuals with a first reinfection in the Omicron BA epoch. Long COVID diagnoses also occurred much closer to the index date following both initial or first reinfection in the Omicron BA epoch as compared to earlier Delta and Omicron epochs. The rate of Long COVID diagnoses has been increasing for reinfections in more recent variants. A number of possible explanations exist for these associative findings. One is that there may be a biological explanation where reinfection may be associated with increased risk of post-acute sequelae. This has been suggested in other literature.^20^ Another explanation is that physician diagnosing behavior has changed and physicians are more likely to have adopted use of either the new U09.9 Long COVID diagnosis code or the existing B94.8 code. This work has also not accounted for other factors like the impact of vaccination status or use of outpatient therapeutics like paxlovid. Future analysis could explore a causal relationship between reinfection and Long COVID and account for these other factors.

A limitation of this study is the reliance on EHR data. EHR limitations are well documented and include selection bias based on varying rates of healthcare utilization, concerns about fitness for purpose and drawing inappropriate conclusions, and data quality and missing data challenges.^43^ EHR studies also differ from clinical studies, where patients may be followed more closely. A major limitation of this particular analysis is that we are limited to EHR collected at specific hospitals; we cannot join patient records between hospitals. Strategies we have used to address these limitations include hospitalization inclusion criteria for sites and visit inclusion criteria for individuals that promote more detailed, robust, and higher quality data.

A second limitation is that it is not feasible to include results of home COVID-19 tests. Individuals may be testing positive for COVID-19 reinfections that are not documented in this dataset. This may result in an underestimate of the number of individuals with reinfections. The varying availability of home tests over the duration considered for this project may result in an uneven impact of this limitation. To address this limitation, we have attempted to maintain a focus on behaviors that require healthcare interaction, such as Long COVID diagnosis, biomarkers, and hospitalization. We do not suggest generalizability of results to situations that would not involve healthcare settings. We also limited analysis to individuals who had a COVID diagnosis prior to May 1, 2022, in an effort to focus on a window in time when home tests were less common. Future work could explore which time periods best capture testing for reinfection.

Reinfections are well documented in an EHR-based cohort from the RECOVER initiative and align with overall incidence rates in the literature. This work validates existing characterization of reinfection as most common in the Omicron epoch and contributes a novel characterization of lower albumin levels after initial COVID-19 infection and leading up to reinfection. The severity of reinfection appears to be associated with the severity of initial infection, and Long COVID diagnoses appear to occur more often following and closer to the index date of reinfection. We describe a number of opportunities for further research with these findings to better understand COVID-19 reinfections.

## Data Availability

All data used in this study is available through the N3C Enclave to approved users. See https://covid.cd2h.org/for-researchers for instructions on how to access the data.

## ACKNOWLEDGMENTS

Authorship was determined using ICMJE recommendations. The content is solely the responsibility of the authors and does not necessarily represent the official views of the National Institutes of Health or N3C or RECOVER.

This study is part of the NIH Researching COVID to Enhance Recovery (RECOVER) Initiative, which seeks to understand, treat, and prevent the post-acute sequelae of SARS-CoV-2 infection (PASC). For more information on RECOVER, visit https://recovercovid.org/. This research was funded by National Institutes of Health (NIH) Agreement OTA OT2HL161847 as part of the Researching COVID to Enhance Recovery (RECOVER) research program. The Data Use Request (DUR) ID is DUR-94BBC49.

We would like to thank the National Community Engagement Group (NCEG), the Patient-Led Research Collaborative (PLRC); all patient, caregiver, and community Representatives; and all the participants enrolled in the RECOVER Initiative. We would like to specifically thank Dr. David Sahner and Dr. Lorna Thorpe for their feedback on the manuscript.

## N3C Attribution

The analyses described in this publication were conducted with data or tools accessed through the NCATS N3C Data Enclave covid.cd2h.org/enclave and supported by CD2H - The National COVID Cohort Collaborative (N3C) IDeA CTR Collaboration 3U24TR002306-04S2 NCATS U24 TR002306. This research was possible because of the patients whose information is included within the data from participating organizations (covid.cd2h.org/dtas) and the organizations and scientists (covid.cd2h.org/duas) who have contributed to the on-going development of this community resource (cite this https://doi.org/10.1093/jamia/ocaa196).

## Institutional Review Board

The N3C data transfer to NCATS is performed under a Johns Hopkins University Reliance Protocol # IRB00249128 or individual site agreements with NIH. The N3C Data Enclave is managed under the authority of the NIH; information can be found at https://ncats.nih.gov/n3c/resources.

## We gratefully acknowledge the following core contributors to N3C

Adam B. Wilcox, Adam M. Lee, Alexis Graves, Alfred (Jerrod) Anzalone, Amin Manna, Amit Saha, Amy Olex, Andrea Zhou, Andrew E. Williams, Andrew Southerland, Andrew T. Girvin, Anita Walden, Anjali A. Sharathkumar, Benjamin Amor, Benjamin Bates, Brian Hendricks, Brijesh Patel, Caleb Alexander, Carolyn Bramante, Cavin Ward-Caviness, Charisse Madlock-Brown, Christine Suver, Christopher Chute, Christopher Dillon, Chunlei Wu, Clare Schmitt, Cliff Takemoto, Dan Housman, Davera Gabriel, David A. Eichmann, Diego Mazzotti, Don Brown, Eilis Boudreau, Elaine Hill, Elizabeth Zampino, Emily Carlson Marti, Emily R. Pfaff, Evan French, Farrukh M Koraishy, Federico Mariona, Fred Prior, George Sokos, Greg Martin, Harold Lehmann, Heidi Spratt, Hemalkumar Mehta, Hongfang Liu, Hythem Sidky, J.W. Awori Hayanga, Jami Pincavitch, Jaylyn Clark, Jeremy Richard Harper, Jessica Islam, Jin Ge, Joel Gagnier, Joel H. Saltz, Johanna Loomba, John Buse, Jomol Mathew, Joni L. Rutter, Julie A. McMurry, Justin Guinney, Justin Starren, Karen Crowley, Katie Rebecca Bradwell, Kellie M. Walters, Ken Wilkins, Kenneth R. Gersing, Kenrick Dwain Cato, Kimberly Murray, Kristin Kostka, Lavance Northington, Lee Allan Pyles, Leonie Misquitta, Lesley Cottrell, Lili Portilla, Mariam Deacy, Mark M. Bissell, Marshall Clark, Mary Emmett, Mary Morrison Saltz, Matvey B. Palchuk, Melissa A. Haendel, Meredith Adams, Meredith Temple-O’Connor, Michael G. Kurilla, Michele Morris, Nabeel Qureshi, Nasia Safdar, Nicole Garbarini, Noha Sharafeldin, Ofer Sadan, Patricia A. Francis, Penny Wung Burgoon, Peter Robinson, Philip R.O. Payne, Rafael Fuentes, Randeep Jawa, Rebecca Erwin-Cohen, Rena Patel, Richard A. Moffitt, Richard L. Zhu, Rishi Kamaleswaran, Robert Hurley, Robert T. Miller, Saiju Pyarajan, Sam G. Michael, Samuel Bozzette, Sandeep Mallipattu, Satyanarayana Vedula, Scott Chapman, Shawn T. O’Neil, Soko Setoguchi, Stephanie S. Hong, Steve Johnson, Tellen D. Bennett, Tiffany Callahan, Umit Topaloglu, Usman Sheikh, Valery Gordon, Vignesh Subbian, Warren A. Kibbe, Wendy Hernandez, Will Beasley, Will Cooper, William Hillegass, Xiaohan Tanner Zhang. Details of contributions available at covid.cd2h.org/core-contributors

## Data Partners with Released Data

### Institutions whose data are released or pending

#### Available

Advocate Health Care Network — UL1TR002389: The Institute for Translational Medicine (ITM) • Boston University Medical Campus — UL1TR001430: Boston University Clinical and Translational Science Institute • Brown University — U54GM115677: Advance Clinical Translational Research (Advance-CTR) • Carilion Clinic — UL1TR003015: iTHRIV Integrated Translational health Research Institute of Virginia • Charleston Area Medical Center U54GM104942: West Virginia Clinical and Translational Science Institute (WVCTSI) • Children’s Hospital Colorado — UL1TR002535: Colorado Clinical and Translational Sciences Institute • Columbia University Irving Medical Center — UL1TR001873: Irving Institute for Clinical and Translational Research • Duke University — UL1TR002553: Duke Clinical and Translational Science Institute • George Washington Children’s Research Institute — UL1TR001876: Clinical and Translational Science Institute at Children’s National (CTSA-CN) • George Washington University — UL1TR001876: Clinical and Translational Science Institute at Children’s National (CTSA-CN) • Indiana University School of Medicine — UL1TR002529: Indiana Clinical and Translational Science Institute • Johns Hopkins University — UL1TR003098: Johns Hopkins Institute for Clinical and Translational Research • Loyola Medicine — Loyola University Medical Center • Loyola University Medical Center — UL1TR002389: The Institute for Translational Medicine (ITM) • Maine Medical Center — U54GM115516: Northern New England Clinical & Translational Research (NNE-CTR) Network • Massachusetts General Brigham — UL1TR002541: Harvard Catalyst • Mayo Clinic Rochester UL1TR002377: Mayo Clinic Center for Clinical and Translational Science (CCaTS) • Medical University of South Carolina — UL1TR001450: South Carolina Clinical & Translational Research Institute (SCTR) • Montefiore Medical Center — UL1TR002556: Institute for Clinical and Translational Research at Einstein and Montefiore • Nemours — U54GM104941: Delaware CTR ACCEL Program • NorthShore University HealthSystem — UL1TR002389: The Institute for Translational Medicine (ITM) • Northwestern University at Chicago — UL1TR001422: Northwestern University Clinical and Translational Science Institute (NUCATS) • OCHIN — INV-018455: Bill and Melinda Gates Foundation grant to Sage Bionetworks • Oregon Health & Science University — UL1TR002369: Oregon Clinical and Translational Research Institute • Penn State Health Milton S. Hershey Medical Center — UL1TR002014: Penn State Clinical and Translational Science Institute • Rush University Medical Center — UL1TR002389: The Institute for Translational Medicine (ITM) • Rutgers, The State University of New Jersey — UL1TR003017: New Jersey Alliance for Clinical and Translational Science • Stony Brook University — U24TR002306 • The Ohio State University — UL1TR002733: Center for Clinical and Translational Science • The State University of New York at Buffalo — UL1TR001412: Clinical and Translational Science Institute • The University of Chicago — UL1TR002389: The Institute for Translational Medicine (ITM) • The University of Iowa — UL1TR002537: Institute for Clinical and Translational Science • The University of Miami Leonard M. Miller School of Medicine — UL1TR002736: University of Miami Clinical and Translational Science Institute • The University of Michigan at Ann Arbor — UL1TR002240: Michigan Institute for Clinical and Health Research • The University of Texas Health Science Center at Houston — UL1TR003167: Center for Clinical and Translational Sciences (CCTS) • The University of Texas Medical Branch at Galveston — UL1TR001439: The Institute for Translational Sciences • The University of Utah UL1TR002538: Uhealth Center for Clinical and Translational Science • Tufts Medical Center UL1TR002544: Tufts Clinical and Translational Science Institute • Tulane University — UL1TR003096: Center for Clinical and Translational Science • University Medical Center New Orleans — U54GM104940: Louisiana Clinical and Translational Science (LA CaTS) Center • University of Alabama at Birmingham — UL1TR003096: Center for Clinical and Translational Science • University of Arkansas for Medical Sciences — UL1TR003107: UAMS Translational Research Institute • University of Cincinnati — UL1TR001425: Center for Clinical and Translational Science and Training • University of Colorado Denver, Anschutz Medical Campus UL1TR002535: Colorado Clinical and Translational Sciences Institute • University of Illinois at Chicago — UL1TR002003: UIC Center for Clinical and Translational Science • University of Kansas Medical Center — UL1TR002366: Frontiers: University of Kansas Clinical and Translational Science Institute • University of Kentucky — UL1TR001998: UK Center for Clinical and Translational Science • University of Massachusetts Medical School Worcester — UL1TR001453: The UMass Center for Clinical and Translational Science (UMCCTS) • University of Minnesota — UL1TR002494: Clinical and Translational Science Institute • University of Mississippi Medical Center — U54GM115428: Mississippi Center for Clinical and Translational Research (CCTR) • University of Nebraska Medical Center — U54GM115458: Great Plains IDeA-Clinical & Translational Research • University of North Carolina at Chapel Hill UL1TR002489: North Carolina Translational and Clinical Science Institute • University of Oklahoma Health Sciences Center — U54GM104938: Oklahoma Clinical and Translational Science Institute (OCTSI) • University of Rochester — UL1TR002001: UR Clinical & Translational Science Institute • University of Southern California — UL1TR001855: The Southern California Clinical and Translational Science Institute (SC CTSI) • University of Vermont — U54GM115516: Northern New England Clinical & Translational Research (NNE-CTR) Network • University of Virginia — UL1TR003015: iTHRIV Integrated Translational health Research Institute of Virginia • University of Washington — UL1TR002319: Institute of Translational Health Sciences • University of Wisconsin-Madison — UL1TR002373: UW Institute for Clinical and Translational Research • Vanderbilt University Medical Center — UL1TR002243: Vanderbilt Institute for Clinical and Translational Research • Virginia Commonwealth University — UL1TR002649: C. Kenneth and Dianne Wright Center for Clinical and Translational Research • Wake Forest University Health Sciences — UL1TR001420: Wake Forest Clinical and Translational Science Institute • Washington University in St. Louis — UL1TR002345: Institute of Clinical and Translational Sciences • Weill Medical College of Cornell University — UL1TR002384: Weill Cornell Medicine Clinical and Translational Science Center • West Virginia University — U54GM104942: West Virginia Clinical and Translational Science Institute (WVCTSI)

## Submitted

Icahn School of Medicine at Mount Sinai — UL1TR001433: ConduITS Institute for Translational Sciences • The University of Texas Health Science Center at Tyler — UL1TR003167: Center for Clinical and Translational Sciences (CCTS) • University of California, Davis — UL1TR001860: UCDavis Health Clinical and Translational Science Center • University of California, Irvine — UL1TR001414: The UC Irvine Institute for Clinical and Translational Science (ICTS) • University of California, Los Angeles — UL1TR001881: UCLA Clinical Translational Science Institute • University of California, San Diego — UL1TR001442: Altman Clinical and Translational Research Institute • University of California, San Francisco — UL1TR001872: UCSF Clinical and Translational Science Institute Pending: Arkansas Children’s Hospital — UL1TR003107: UAMS Translational Research Institute • Baylor College of Medicine — None (Voluntary) • Children’s Hospital of Philadelphia UL1TR001878: Institute for Translational Medicine and Therapeutics • Cincinnati Children’s Hospital Medical Center — UL1TR001425: Center for Clinical and Translational Science and Training • Emory University — UL1TR002378: Georgia Clinical and Translational Science Alliance • HonorHealth — None (Voluntary) • Loyola University Chicago — UL1TR002389: The Institute for Translational Medicine (ITM) • Medical College of Wisconsin — UL1TR001436: Clinical and Translational Science Institute of Southeast Wisconsin • MedStar Health Research Institute — UL1TR001409: The Georgetown-Howard Universities Center for Clinical and Translational Science (GHUCCTS) • MetroHealth — None (Voluntary) • Montana State University — U54GM115371: American Indian/Alaska Native CTR • NYU Langone Medical Center — UL1TR001445: Langone Health’s Clinical and Translational Science Institute • Ochsner Medical Center — U54GM104940: Louisiana Clinical and Translational Science (LA CaTS) Center • Regenstrief Institute — UL1TR002529: Indiana Clinical and Translational Science Institute • Sanford Research — None (Voluntary) • Stanford University — UL1TR003142: Spectrum: The Stanford Center for Clinical and Translational Research and Education • The Rockefeller University — UL1TR001866: Center for Clinical and Translational Science • The Scripps Research Institute — UL1TR002550: Scripps Research Translational Institute • University of Florida — UL1TR001427: UF Clinical and Translational Science Institute University of New Mexico Health Sciences Center — UL1TR001449: University of New Mexico Clinical and Translational Science Center • University of Texas Health Science Center at San Antonio — UL1TR002645: Institute for Integration of Medicine and Science • Yale New Haven Hospital — UL1TR001863: Yale Center for Clinical Investigation

