## Supplemental Tables and Figures for "SARS-CoV-2 Reinfection is Preceded by Unique Biomarkers and Related to Initial Infection Timing and Severity: an N3C RECOVER EHR-Based Cohort Study"

**Supplementary Information**

Supplementary Table 1. Descriptive characteristics of multiple reinfected and non-reinfected COVID-19 positive patients

| **Category** | **Variable** | **No Reinfection** (N=1505855) | **One Reinfection** (N=89357) | **Two Reinfection** (N=2176) | **Three or More Reinfection** (N=102) | **Total** (N=1597490) | **P value** |
|---|---|---|---|---|---|---|---|
| **Age, Mean (SD)** | Age | 49.14 (18.26) | 44.29 (17.69) | 44.73 (18.48) | 48.23 (18.07) | 48.86 (18.27) | <0.0001 |
| **Sex N (%)** | Female | 946166 (62.83) | 60426 (67.62) | 1500 (68.93) | 63 (61.76) | 1008155 (63.11) | |
| | Male | 559280 (37.14) | 28903 (32.35) | 675 (31.02) | 39 (38.24) | 588897 (36.86) | <0.0001 |
| | No sex Information | 409 (0.03) | 28 (0.03) | <20 | 0 (0) | 438 (0.03) | |
| **Race N (%)** | White | 1141563 (75.81) | 68129 (76.24) | 1641 (75.41) | 82 (80.39) | 1211415 (75.83) | |
| | Black | 199891 (13.27) | 12547 (14.04) | 315 (14.48) | <20 | 212762 (13.32) | |
| | Asian | 24615 (1.63) | 1000 (1.12) | 22 (1.01) | 0 (0) | 25637 (1.6) | |
| | Others | 33996 (2.26) | 2166 (2.42) | 53 (2.44) | <20 | 36218 (2.27) | <0.0001 |
| | No race Information | 105790 (7.03) | 5515 (6.17) | 145 (6.66) | <20 | 111458 (6.98) | |
| **Ethnicity N (%)** | Not Hispanic | 1197960 (79.55) | 71894 (80.46) | 1725 (79.27) | 87 (85.29) | 1271666 (79.6) | |
| | Hispanic | 106956 (7.1) | 7678 (8.59) | 240 (11.03) | <20 | 114881 (7.19) | <0.0001 |
| | No Ethnicity information | 200939 (13.34) | 9785 (10.95) | 211 (9.7) | <20 | 210943 (13.2) | |

Supplementary Table 2. Descriptive characteristics of multiple reinfected and non-reinfected COVID-19 positive patients by hospitalization status

| **Category** | **Variable** | **Not reinfected not hospitalized** (N=1,407,508) | **Not reinfected hospitalized** (N=98,347) | **Reinfected not hospitalized** (N=86,027) | **Reinfected hospitalized** (N=5,608) | **Total** (N=1,597,490) | **P value** |
|---|---|---|---|---|---|---|---|
| **Age, Mean (SD)** | Age | 48.36 (18.03) | 60.3 (17.97) | 43.49 (17.26) | 56.89 (19.64) | 48.86 (18.27) | <0.0001 |
| **Sex N (%)** | Female | 892404 (63.4) | 53762 (54.67) | 58737 (68.28) | 3252 (57.99) | 1008155 (63.11) | |
| | Male | 514698 (36.57) | 44582 (45.33) | 27261 (31.69) | 2356 (42.01) | 588897 (36.86) | <0.0001S |
| | No sex Information | 406 (0.03) | <20 | 29 (0.03) | 0 (0) | 438 (0.03) | |
| **Race N (%)** | White | 1075594 (76.42) | 65969 (67.08) | 66109 (76.85) | 3743 (66.74) | 1211415 (75.83) | |
| | Black | 178439 (12.68) | 21452 (21.81) | 11655 (13.55) | 1216 (21.68) | 212762 (13.32) | |
| | Asian | 22975 (1.63) | 1640 (1.67) | 932 (1.08) | 90 (1.6) | 25637 (1.6) | |
| | Others | 32151 (2.28) | 1845 (1.88) | 2133 (2.48) | 89 (1.59) | 36218 (2.27) | <0.0001 |
| | No race Information | 98349 (6.99) | 7441 (7.57) | 5198 (6.04) | 470 (8.38) | 111458 (6.98) | |
| **Ethnicity (N, %)** | Not Hispanic | 1119820 (79.56) | 78140 (79.45) | 69422 (80.7) | 4284 (76.39) | 1271666 (79.6) | |
| | Hispanic | 98271 (6.98) | 8685 (8.83) | 7313 (8.5) | 612 (10.91) | 114881 (7.19) | <0.0001 |
| | No Ethnicity information | 189417 (13.46) | 11522 (11.72) | 9292 (10.8) | 712 (12.7) | 210943 (13.2) | |

Supplementary Figure 1. Filtering process with sex ratio.

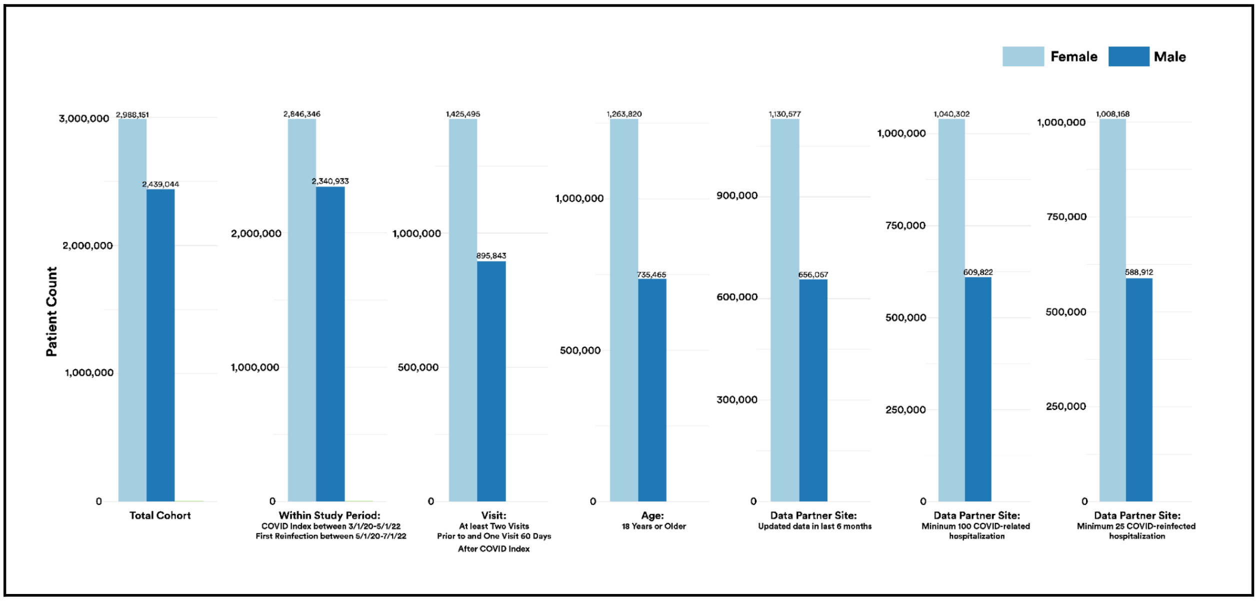

Supplementary Table 3: Count of Severity of Reinfection versus Severity of Initial Infection, Disaggregated by Age

| | | **Severity of Reinfection** | | | | | |
|---|---|---|---|---|---|---|---|
| **Severity of Initial Infection** | **Age** | *No Reinfection* | *Mild with No ED Visit or Hospitalization Around Reinfection Index* | *Mild with ED Visit Around Reinfection Index* | *Moderate with Hospitalization Around Reinfection Index* | *Severe with ECMO or IMV or Vasopressor During Hospitalization Around Reinfection Index* | *Death Within 60 Days of Reinfection Index* |
| *Mild with No ED Visit or Hospitalization Around COVID Index* | 18 - 50 | 701034 | 48776 | 3377 | 1108 | 201 | 64 |
| | 51 - 64 | 340769 | 14559 | 991 | 797 | 94 | 147 |
| | 65+ | 246989 | 7791 | 602 | 1056 | 119 | 411 |
| *Mild with ED Visit Around COVID Index* | 18 - 50 | 64661 | 2957 | 1380 | 198 | 46 | <20 |
| | 51 - 64 | 29429 | 901 | 289 | 102 | <20 | <20 |
| | 65+ | 21650 | 499 | 151 | 132 | <20 | 22 |
| *Moderate with Hospitalization Around COVID Index* | 18 - 50 | 23189 | 911 | 206 | 333 | 37 | <20 |
| | 51 - 64 | 25542 | 750 | 175 | 286 | 35 | 46 |
| | 65+ | 37228 | 835 | 142 | 415 | 52 | 114 |
| *Severe with ECMO or IMV or Vasopressor During Hospitalization Around COVID Index* | 18 - 50 | 3425 | 117 | <20 | 24 | <20 | <20 |
| | 51 - 64 | 2869 | 72 | <20 | 42 | <20 | <20 |
| | 65+ | 3063 | 62 | <20 | 43 | <20 | <20 |
| *Death Within 60 Days of COVID Index* | 18 - 50 | 431 | NA | NA | NA | NA | NA |
| | 51 - 64 | 1352 | NA | NA | NA | NA | NA |
| | 65+ | 4224 | NA | NA | NA | NA | NA |
